# Using Artificial Intelligence-based models to predict the risk of Mucormycosis among COVID-19 Survivors: An Experience from India

**DOI:** 10.1101/2021.09.13.21263511

**Authors:** Shabbir Syed-Abdul, A. Shoban Babu, Raja Shekhar Bellamkonda, Ramaiah Itumalla, GVRK Acharyulu, Surya Krishnamurthy, Y. Venkat Santosh Ramana, Naresh Mogilicharla, Shwetambara Malwade, Yu-Chuan (Jack) Li

**Affiliations:** International Center for Health Information Technology, College of Medical Science and Technology, Taipei Medical University, Taipei, Taiwan; Department of ENT, Gandhi Medical College and Hospital, Secunderabad, Telangana, India; School of Management Studies, University of Hyderabad, Hyderabad, Telangana, India; Department of Health Management, College of Public Health and Health Informatics, University of Hail, Hail, Kingdom of Saudi Arabia; Data Science, iQGateway, Bengaluru, Karnataka, India; Department of Hospital Administration, Gandhi Hospital, Secunderabad, Telangana, India

**Keywords:** artificial intelligence, coronavirus, COVID-19, fungal infection, India, mucormycosis

## Abstract

**Introduction:** India reported a severe public health challenge not only due to the COVID-19 outbreak but also the increasing number of associated mucormycosis cases since 2021. This study aimed at developing artificial intelligence-based models to predict the risk of mucormycosis among the patients at the time of discharge from the hospital.

**Methods:** The dataset included 1229 COVID-19 positive patients, and additional 214 inpatients, COVID-19 positive as well as infected with mucormycosis. We used logistic regression, decision tree, and random forest, and the extreme gradient boosting algorithm. All our models were evaluated with 5-fold validation to derive a reliable estimate of the model error.

**Results:** The logistic regression, XGBoost, and random forest performed equally well with AUROC 95.0, 94.0, and 94.0 respectively. This study also determined the top five variables namely obesity, anosmia, de novo diabetes, myalgia, and nasal discharge, which showed a positive impact on the risk of mucormycosis.

**Conclusion:** The developed model has the potential to predict the patients at high risk and thus, consequently initiating preventive care or aiding in early detection of mucormycosis infection. Thus, this study holds potential for early treatment and better management of patients suffering from COVID-19 associated mucormycosis.

## Introduction

Mucormycosis (previously called zygomycosis) is a life-threatening but rare fungal infection caused by a group of molds called mucormycetes [1]. It is an opportunistic infection characterized by sinusitis and orbital cellulitis that penetrates the arteries to produce thrombosis of the ophthalmic and internal carotid arteries, and later invades the veins and lymphatics [2]. India, the second most populated country in the world has been facing public health challenge due to the outbreak of the COVID-19 [3]. As of July 8th 2021, in India 4,60,704 active COVID-19 cases and 4,05,028 deaths were reported [4]. At this crucial juncture, increasing number of the mucormycosis cases in country are alarming the policy makers and healthcare experts emphasizing the great necessity of preventive measures to detect and treat the patients with mucormycosis. Until 28^th^ June 2021, India reported 40,845 cases of mucormycosis, and considering hundreds of thousands of COVID-19 active cases this number is expected to increase significantly. As of 21^st^ July, 2021, India recorded more than 4300 deaths from COVID-19 related mucormycosis [5]. Difficult diagnosis and relapse of the disease are among the factors hampering treatment success. Therefore, this study is aimed to develop artificial intelligence (AI)-based models to predict the risk of mucormycosis among the COVID-19 patients at the time of their discharge from hospital.

## Methods

### Data pre-processing

This single-center retrospective study was conducted at a tertiary public hospital, designated for COVID-19 and mucormycosis treatment at the Telangana state in India. The dataset comprised of 1229 COVID-19 positive patients, and additional 214 inpatients, those who were tested positive for COVID-19 and subsequently infected with mucormycosis. All these patients were hospitalized and received treatment between 19th March, 2021 and 30th June 2021. Since the patient group with mucormycosis included only those in the age group between 30 to 75 years, we filtered all the patients in the dataset to that age group to ensure that the models are unbiased. The original dataset from medical records contains 74 features from which relevant variables were filtered and encoded for predictive modeling. The categorical variables were binary/one-hot encoded. After encoding, 35 features were input to our prediction models. The final set of features used for modeling is as below:

- Nominal features:
  - Sex: male, female
  - Place of Residence: urban, rural
  - Duration of diabetes: chronic, de novo, none
  - Covid-19 symptoms: severity, asymptomatic, throat pain, anosmia, nasal obstruction, loss of taste, headache, nasal discharge, myalgia, cough, fever, shortness of breath
  - Comorbidities: dialysis, obesity, lung disease, hypothyroid, heart disease, renal disease, hypertension, diabetes, none
  - Medications: antivirals, mineral supplements, anti-parasitics, blood thinners, steroids, vitamin supplements, broad spectrum antibiotics
- Ordinal features:
  - Covid-19 severity: mild, moderate, severe
- Numerical features:
  - Age: 30 to 75

### Modelling

We used logistic regression, decision tree and random forest from the scikit-learn [6]^5^ library and the extreme gradient boosting algorithm from the XGBoost [7] library. To prevent overfitting and selection bias from our skewed dataset we tuned the models to generalize well on unseen data. Regularization was adapted (L2 for logistic regression and XGBoost), wherever applicable and the model parameters were chosen to reduce the model complexity (limiting the depth of the trees to 5, reducing XGBoost to 100 trees and increasing the forest to 500 trees). We used class weights in all our models to handle the class imbalance. We evaluated all our models with 5-fold validation to derive a reliable estimate of the model error.

## Results

The AUROC, recall, precision and accuracy from the different models used with the 5-fold validation are shown in figure 1. The logistic regression, XGBoost and random forest performed equally well with AUROC 95.0, 94.0, and 94.0 respectively. The best accuracy and precision (PPV) were 0.91 ± 0.026 and 0.67 ± 0.0526, respectively achieved by XGBoost, followed by logistic regression. Figure 2 shows box plot of coefficients for all the features across the 5 folds indicating variables having positive and negative impact on the risk of mucormycosis.

**Figure 1:**
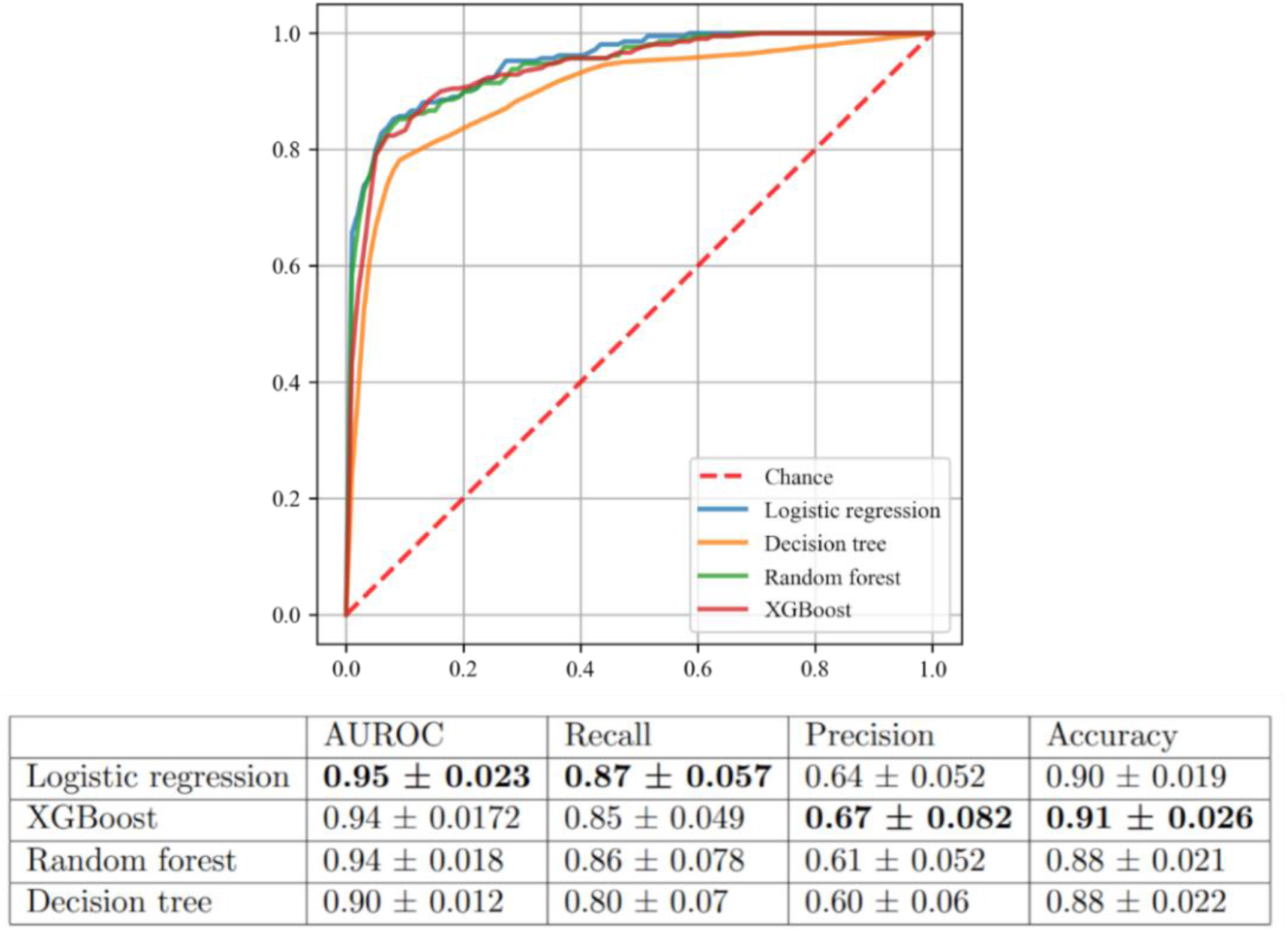
Area under receiver operating characteristic curve (AUROC) for the models developed in the study.

**Figure 2:**
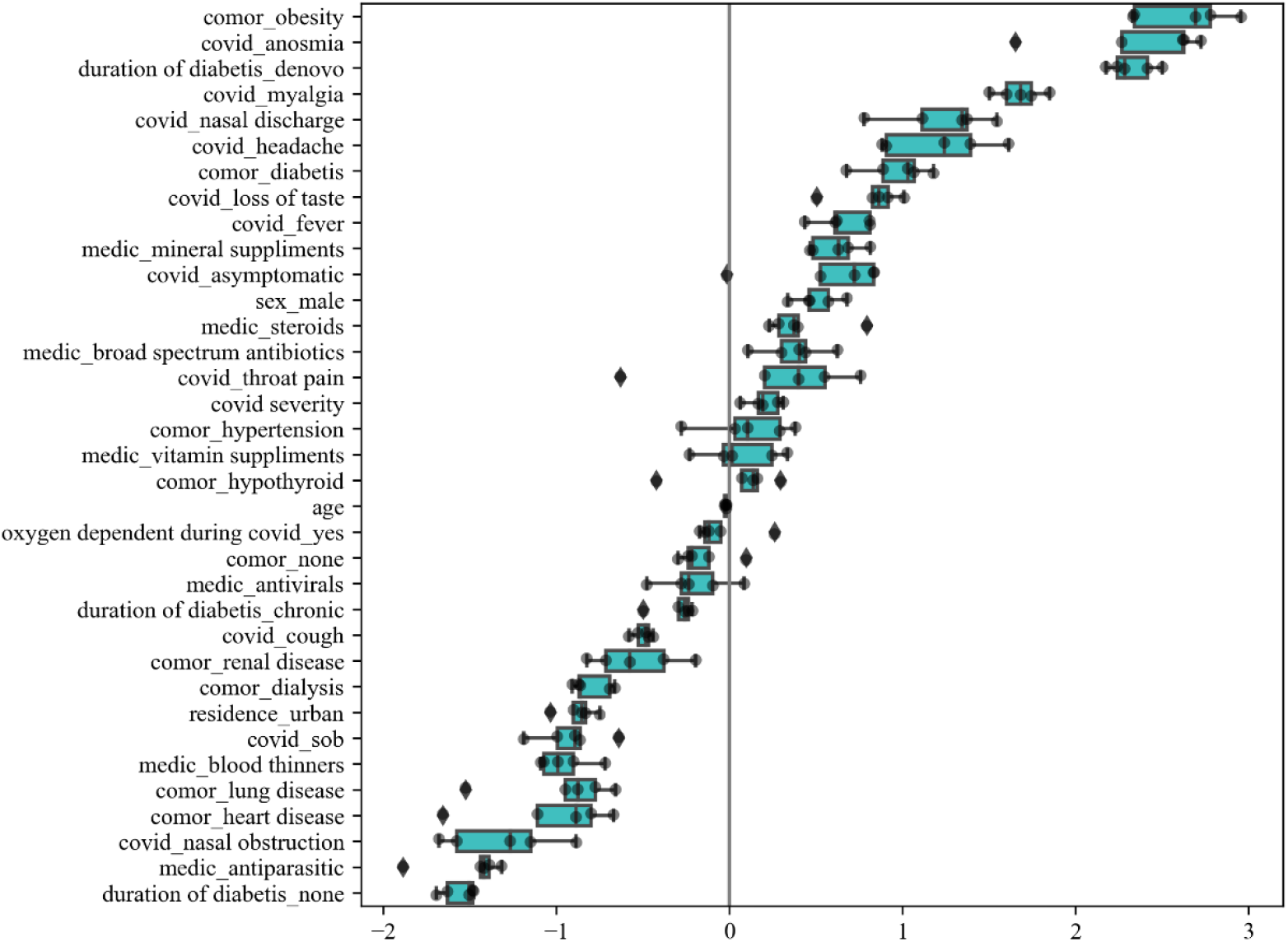
Box plot of coefficients for all the features across the 5 folds showing positive and negative impact on the risk of mucormycosis

## Discussion

To the best of our knowledge, this is the first study to develop an artificial intelligence-based model to predict the risk of mucormycosis among COVID-19 survivors. Our developed models achieved a high AUROC of 0.95± 0.023. Moreover, this study also indicated the probable risk factors having predisposition towards mucormycosis. Features such as obesity, anosmia, de novo diabetes, myalgia, and nasal discharge were the top five variables showed positive impact towards risk of mucormycosis. Previous research by Mahalakshmi et al, Singh et al and Pakdel et al also indicated that uncontrolled diabetes can be a strong pre-disposing factor for post COVID mucormycosis [8] [9] [10]. Some studies also pointed out other risk factors for mucormycosis, such as treatment with corticosteroids, those who underwent organ transplantation, or those suffering from hematological malignancies [11] [8] [12]. Our study was the first one to reveal de novo diabetes obesity, anosmia, myalgia and nasal discharge as the factors associated with development of mucormycosis in COVID-19 patients.

On the other hand, absence of diabetes mellitus, under antiparasitic medications, nasal obstruction were the symptoms negatively associated with the mucormycosis. These models can be used to predict the risk and aid healthcare providers to take preventive care for the patients who are at high risk to develop mucormycosis. Mortality from the disease is relatively high and early diagnosis along with preventive measures would be the key to survival [9]. Our study, indicates potential for early detection, early treatment and better management of patients suffering from COVID-19 associated mucormycosis. The AI models can not only detect high risk patients, but also indicate the possible pre-disposing factors that need to be taken care of post COVID-19 treatment.

However, there are some limitations for generalization of these models, which need further validations with dataset from other states of India. Despite this, the study’s model has the potential to predict the patients at high risk and thus, consequently initiating preventive care or aiding in early detection of mucormycosis infection. Moreover, including a dataset for a longer time span would enable more accurate results with a larger dataset. Future studies could include predicting the number of days after discharge, when a patient has a possibility of developing mucormycosis.

## Data Availability

The datasets generated during and/or analyzed during the current study are available from the corresponding author on reasonable request.

## Funding

This work is supported in part by Ministry of Science and Technology, Taiwan [grant numbers 108-2221-E-038-013, 110-2923-E-038 -001 -MY3]; Taipei Medical University, Taiwan [grant numbers 108-3805-009-110, 109-3800-020-400]; Ministry of Education, Taiwan [grant number 108-6604-002-400]; Wanfang hospital, Taiwan [grant number 106TMU-WFH-01-4].

## Notes

### Competing Interest Statement

The authors have declared no competing interest.

### Author Declarations

This study was approved by the Gandhi Medical College & Hospital Institutional Ethical Committee, Secunderabad, Telangana, India (Rc. No. IEC/GMC/2021, dated 21/June/2021 "Predicting the risk of Mucormycosis among COVID-19 Survivors using Artificial Intelligence based models"). This study was performed in accordance with the Helsinki Declaration of 1964, and its later amendments.

